# Predicting High-Risk Fetal Cardiac Disease Anticipated to Need Immediate Postnatal Stabilization and Intervention with Planned Pediatric Cardiac Operating Room Delivery

**DOI:** 10.1101/2023.03.13.23287237

**Authors:** Amol Moray, Proscovia M. Mugaba, Chloe Joynt, Angela McBrien, Luke Eckersley, Ernest Philipos, Paula Holinski, Lindsay Ryerson, James Y Coe, Sujata Chandra, Amanda Aiken, Billy Wong, Michele Derbyshire, Maria Lefebvre, Mohamed Al Aklabi, Lisa K Hornberger

**Affiliations:** Fetal & Neonatal Cardiology Program, Division of Cardiology, Department of Pediatrics, The Mazankowski Alberta Heart Institute and Women’s & Children’s Health Research Institute, University of Alberta; Division of Neonatology, Department of Pediatrics, The Mazankowski Alberta Heart Institute and Women’s & Children’s Health Research Institute, University of Alberta; Division of Critical Care, The Mazankowski Alberta Heart Institute and Women’s & Children’s Health Research Institute, University of Alberta; Department of Anesthesia, The Mazankowski Alberta Heart Institute and Women’s & Children’s Health Research Institute, University of Alberta; Interventional Cardiology, Division of Cardiology, Department of Pediatrics, The Mazankowski Alberta Heart Institute and Women’s & Children’s Health Research Institute, University of Alberta; Department of Obstetrics & Gynecology, The Mazankowski Alberta Heart Institute and Women’s & Children’s Health Research Institute, University of Alberta; Stollery Pediatric and Mazankowski Cardiac Operating Rooms, The Mazankowski Alberta Heart Institute and Women’s & Children’s Health Research Institute, University of Alberta; Division of Pediatric Cardiovascular Surgery, Department of Surgery; The Mazankowski Alberta Heart Institute and Women’s & Children’s Health Research Institute, University of Alberta

## Abstract

**Background:** Distances between delivery centers and cardiac services can make the care of fetuses with cardiac disease(CD) at risk of acute cardiorespiratory instability(ACRI) at birth a challenge. In 2013 we implemented a fetal echocardiography(FE)-based algorithm targeting fetuses considered high-risk for ACRI at ≤2 hours of birth for Caesarian section(CS) delivery in our pediatric cardiac operating room(PCOR) of our children’s hospital. We examine the experience and outcomes of affected newborns.

**Methods:** We reviewed maternal and postnatal medical records of all fetuses with CD at high-risk for ACRI encountered January 2013-March 2022. Secondary analysis was performed including all fetuses with diagnoses of d-transposition of the great arteries/intact ventricular septum(d-TGA/IVS) and hypoplastic left heart syndrome(HLHS) encountered over the study period.

**Results:** Forty fetuses were considered high-risk for ACRI: 15 d-TGA/IVS and 7 HLHS with restrictive atrial septum(RAS), 4 absent pulmonary valve syndrome, 3 obstructed anomalous pulmonary veins, 2 severe Ebstein anomaly, 2 thoracic/intracardiac tumors and 7 others. PCOR delivery occurred for 33 but not for 7 (5 d-TGA/IVS, 2 HLHS with RAS). For high-risk cases, FE had a positive predictive value of 50% for intervention/ECMO/death at ≤2 hours and 70% at ≤24 hours. Of “low-risk” cases, 6/46 with d-TGA/IVS and 0/45 with HLHS required intervention at ≤2 hours. FE predicted intervention/ECMO/death at ≤2hours with a sensitivity of 67%, specificity 93%, and positive and negative predictive values of 87% and 87%, respectively, for d-TGA/IVS, and 100%, 95%, 71%, and 100% for HLHS, respectively.

**Conclusions:** FE predicts need for urgent intervention in majority with d-TGA/IVS and HLHS, and in half of the entire spectrum of high-risk CD.

## INTRODUCTION

Advances in fetal echocardiography (FE) have made it possible to accurately evaluate fetal cardiac anatomy and circulatory physiology in a significant proportion of fetuses with cardiac disease (CD).^1,2^ This has enabled the prediction of the intrauterine and perinatal course of most fetal CD,^2,3^ which, in turn has led to improved perinatal care and the development of intrauterine interventions preventing the evolution of more severe secondary cardiac pathology.^2-7^

In more recent years, FE has increasingly facilitated recognition of a subset of critical CD with potential for acute cardiorespiratory instability (ACRI) soon after birth. Prenatal predictors of ACRI are evolving, particularly for d-transposition of the great arteries with intact ventricular septum (d-TGA/IVS)^8-11^ and hypoplastic left heart syndrome (HLHS) with a restrictive or intact atrial septum (RAS).^12,13^ Other critical CD lesions have also been recognized as potentially life-threatening at birth.^14-18^ With this knowledge, FE-based algorithms have evolved to guide perinatal management, including mode and location of delivery. Only two existing studies report promising results of how this approach could ultimately improve the clinical outcomes of these challenging patients^3,19^ ; however further data are needed that explore approaches with different types of multidisciplinary infrastructure and geographical challenges faced by many pediatric cardiac programs internationally.

It is not uncommon for obstetrical services to be separated by a short, but important, distance from the pediatric cardiac surgical or interventional centers. This renders the ex-utero transport of a critically ill newborn to the cardiac center a challenge, and may reduce the benefit of a prenatal diagnosis. There has been a move by some centers towards delivery of such fetuses under controlled circumstances in a pediatric cardiac operating room (PCOR) with a full cardiac team who can stabilize the newborn by performing cardiac intervention within minutes of birth.^3,19^ There is a paucity of data on the efficacy of this approach and its risks in the published literature.

In 2013, our Western Canadian centralized pediatric cardiac program initiated a FE-based algorithm using a multi-disciplinary team approach to risk-stratify and triage fetuses for specialized delivery room care, 5.4 kilometers from high-risk obstetrical services. The current study reviews our experience and assesses the accuracy of FE in predicting the need for urgent postnatal intervention for a spectrum of high-risk pathologies. We further focus on the more common lesions of d-TGA/IVS and HLHS with RAS to examine the sensitivity and specificity of current prenatal predictors.

## METHODS

This retrospective cohort study was approved of by the Human Research Ethics Committee of the University of Alberta. Pregnancies complicated by major structural and functional fetal CD, including life-threatening arrhythmias, evaluated and managed in our fetal and neonatal cardiology program from January 2013 to March 2022 were identified through our institutional fetal cardiology database. Firstly, those complicated by fetal CD assigned to delivery room level of care (LOC) 4 as per the American Heart Association Fetal Cardiology Guidelines^20^ were analyzed. We then performed analysed all pregnancies complicated by a fetal diagnosis of d-TGA/IVS and HLHS evaluated and delivered at our institution over the time same period.

### Fetal Echocardiography-Based Algorithm

In 2013, given the distance between our quaternary obstetrical hospital, the Lois Hole Hospital for Women, and the pediatric cardiac program at the Stollery Children’s Hospital, our institution evolved a multi-disciplinary team approach guided by FE to risk-stratify CD^20^: infants having non-ductus arteriosus dependent CD remained at the obstetrical hospital (LOC 1-2), those with ductus arteriosus-dependent CD were transferred to the Stollery Children’s Hospital within hours of birth (LOC 2-3), and CD cases deemed at high-risk of ACRI were delivered in our PCOR by Caesarian section (CS) (LOC 4). FE evaluation in each case entailed assessing for lesion-specific criteria to evaluate CD severity using published data, whenever available, and/or clinical experience with multidisciplinary team consensus. In d-TGA/IVS, we assessed for RAS and a small/restrictive ductus arteriosus,^8-11^ which include tethered, fixed position of septum primum, small atrial flow orifice (≤3mm), ductus arteriosus constriction, and increased ductal flow velocities suggestive of obstruction. Serial studies were performed to assess for evolving features with ≥2 exams after 35 weeks for most. In HLHS, we assessed for features of RAS including foramen ovale size <3mm, thick atrial septum and pulmonary vein Doppler flow forward/reverse velocity-time integral (FR VTI) ratio, with a ratio of <3 near term suggestive of severe left atrial hypertension necessitating emergent intervention.^12^ Maternal hyperoxygenation was performed as previously described.^21^

For other prenatally diagnosed CD at risk of ACRI, we assessed for general and lesion-specific features as elaborated in Table 1, including presence/absence of hydrops, cardiac size, combined cardiac output, and evidence of biventricular dysfunction as previously reported.^2-4,14-18^ Where available, noncardiac data was acquired through ultrasound and fetal magnetic resonance imaging for extracardiac pathology and features of airway obstruction (TOF/APV)^16^ or lung parenchyma suggestive of ‘nutmeg lung’ (HLHS with RAS and obstructed total anomalous pulmonary venous drainage (TAPVD)).^22^

**Table 1.**
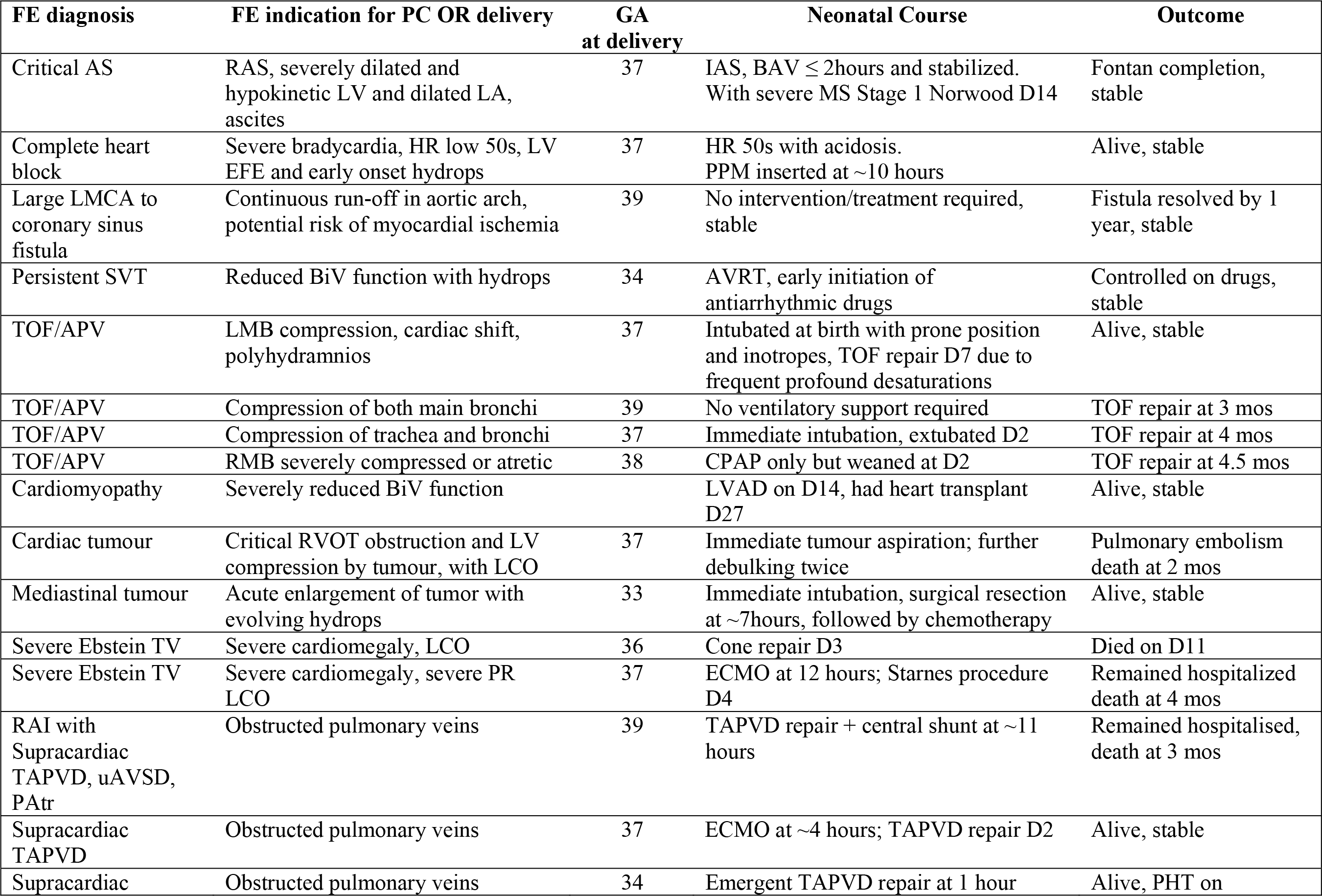
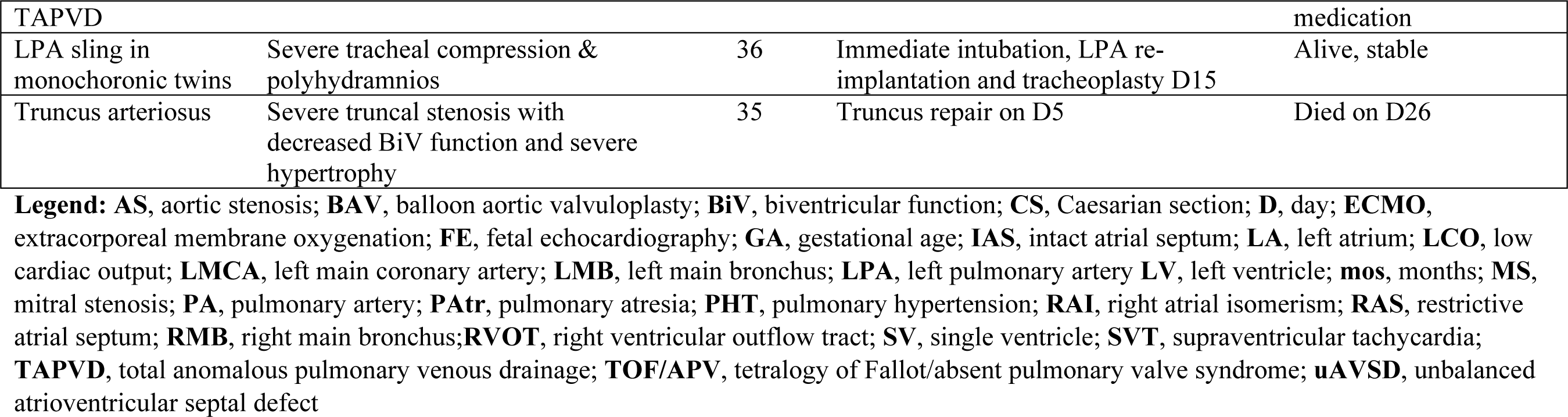
Fetal Cardiac Pathology Recommended for Delivery in the Pediatric Cardiac Operating Room.

### Pediatric Cardiac Operating Room Delivery Planning

For those fetuses deemed at highest-risk of ACRI (LOC 4), expected to require an intervention at ≤2 hours and/or considered at exceptionally high-risk during inter-hospital transport, CS delivery in the PCOR was recommended with the cardiac team on standby to carry out acute neonatal care. CS delivery was based on the predicted need for urgent postnatal interventions dependent on a pre-assembled multidisciplinary team with specific technical skills. For delivery in the PCOR, the fetus had to have no major extracardiac pathology including chromosomal/genetic anomaly that would make aggressive management unlikely to alter a poor prognosis. The mother could not be deemed at high-risk given the lack of obstetrical support in the PC hospital. Our management algorithm is presented in Figure 1.

**Figure 1.**
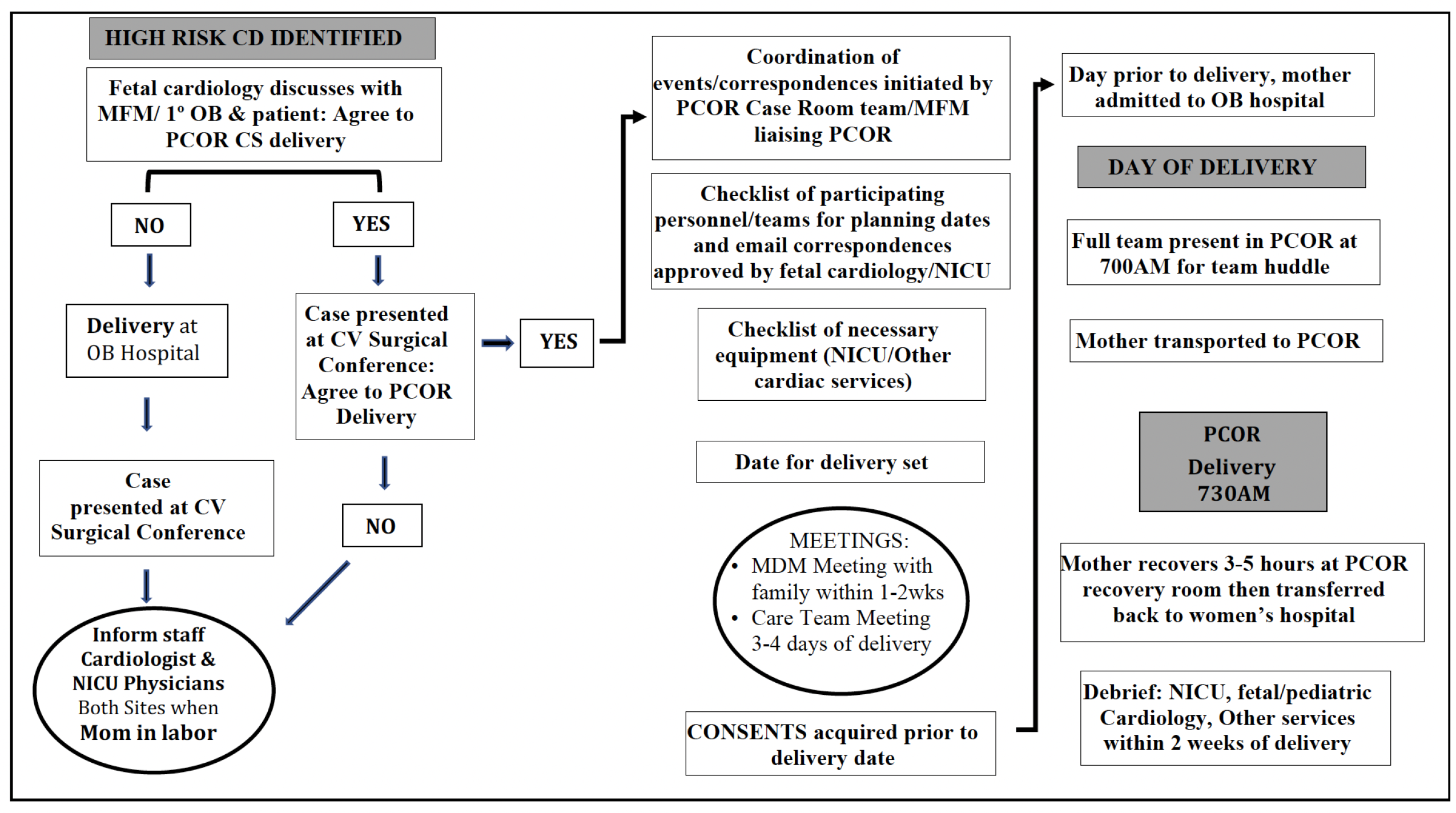
Algorithm for High Risk Fetal Cardiac Disease with Delivery in the PCOR. CD-cardiac disease; CV-cardiovascular; MFM/1ºOB-maternal-fetal medicine specialist/primary obstetrician; NICU-neonatal intensive care unit/service; OB-obstetrical; OR-operating room; PCOR-pediatric cardiac operating room.

Once a pregnancy was identified as potentially meeting criteria for PCOR delivery, and the patient agreed, clinical details were reviewed by a multidisciplinary team, and, with the group’s consensus, the delivery plan was made, taking into consideration the ideal gestational age, date, needs at delivery for acute neonatal care and disposition of the newborn (surgical neonatal intensive care unit versus pediatric cardiac intensive care unit) depending on likelihood for needing extracorporeal membrane oxygenation (ECMO). The teams involved in the consensus included obstetrical, neonatology, and fetal cardiology specialists consistently and other pediatric cardiology subspecialists as applicable, cardiac surgeons, pediatric cardiac critical care, pediatric cardiac anesthesia and both obstetrical and neonatal nursing. Delivery was planned for 38-39 weeks of gestation unless there was a high-risk of earlier delivery or acute fetal compromise/death. On the morning of delivery, both maternal and neonatal multidisciplinary teams assembled prior to the delivery in preparation for enacting the perinatal care plan. Delivery occurred in one PCOR room and the baby was transported at birth to an adjoining PCOR where the cardiac care team was available for resuscitation. Following CS, the mother was monitored by an obstetrical nurse for 3-5 hours in the recovery room to ensure clinical stability prior to being transported back to the obstetrical hospital for further care. It was understood, should the mother go into spontaneous labor, delivery would occur in the obstetrical hospital with early notification of the multidisciplinary team and urgent neonatal transport protocols enacted.

### Data Collection and Analysis

Demographic and clinical data were obtained through a chart review of fetal/postnatal echocardiograms and medical records. We extracted data on the fetal CD and findings suggesting risk of ACRI, delivery room and postnatal care plan, delivery information, postnatal findings and management and their timing from birth, and postnatal outcomes. For those in whom CS in the PCOR occurred, we analyzed maternal postnatal outcomes documented in the same admission. We compared the fetal diagnosis and delivery room and postnatal care plans with the postnatal diagnosis, clinical course and treatment/interventions performed and we used this to assess the utility of using FE in identifying high-risk fetal CD. We further defined its accuracy in predicting postnatal ACRI (positive predictive value), for purposes of this review defined as a combined outcome of intervention/ECMO/death occurring at ≤2 hours and ≤24 hours after birth. We performed a sub-analysis of all fetal d-TGA/IVS and HLHS diagnosed prenatally in our program and managed at the Stollery during the study period. We determined sensitivity, specificity, positive and negative predictive values of FE for fetuses at risk of postnatal ACRI shortly after birth.

## RESULTS

Over the study period, 670 fetuses with CD or persistent brady or tachyarrhythmias were encountered in our program. Of these, 40(6.1%) were identified for delivery in the PCOR, including 37 singleton and 3 twin pregnancies (one affected twin each). Figure 2 summarizes the CD diagnoses and outcomes of this cohort, all confirmed to have no major discrepancies between the prenatal and postnatal cardiac diagnoses after birth. The largest CD subgroups included 15 with d-TGA/IVS and 7 with HLHS with RAS. The 18 others included 4 with TOF/APV, 3 with TAPVD with obstruction, 2 with severe Ebstein anomaly, 2 with persistent, life-threatening arrhythmias and 7 with other CD (Table 1). Figures 3-5 demonstrate examples of ACRI cases.

**Figure 2.**
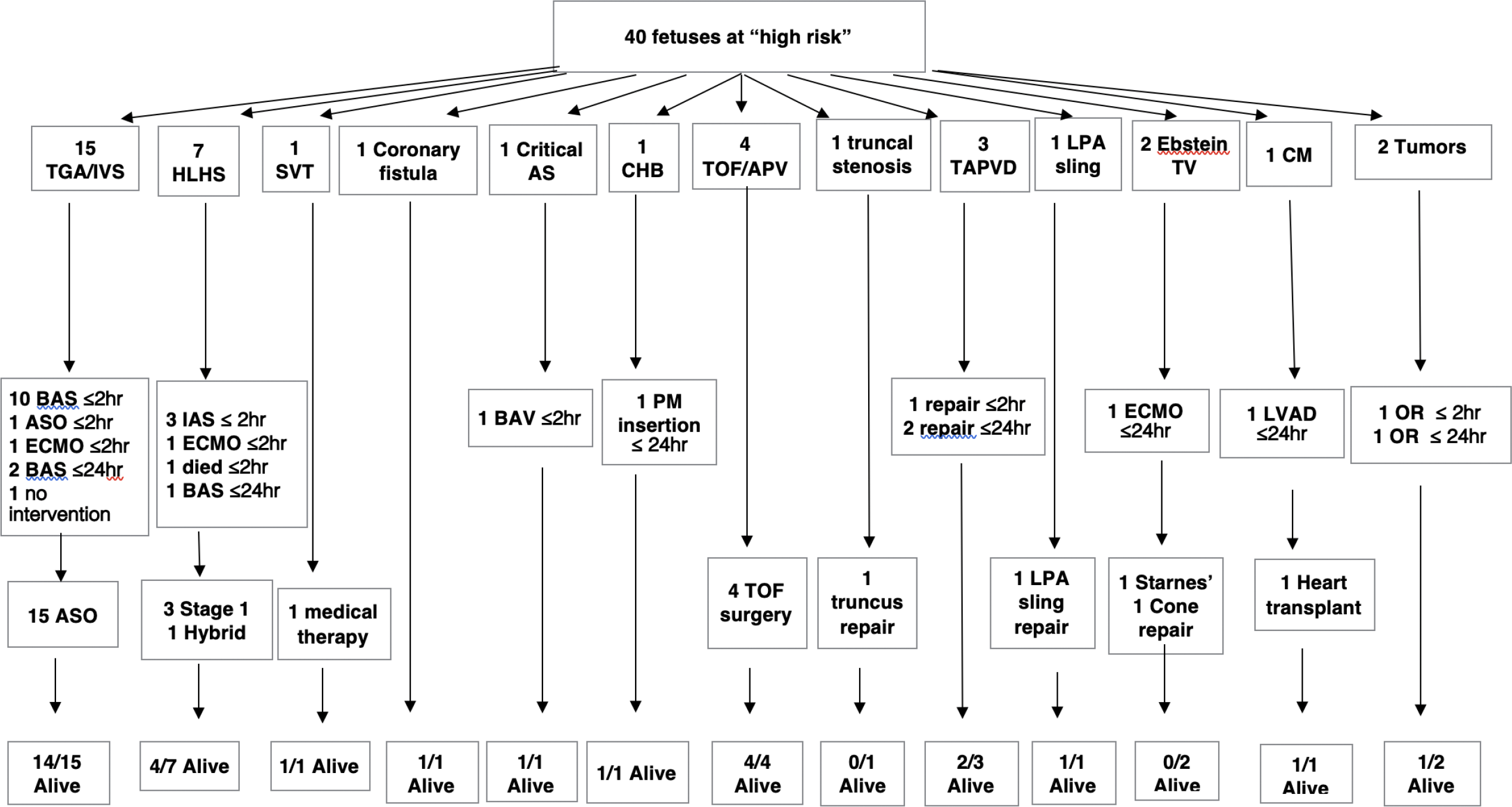
Outcomes of the Fetuses Considered at High Risk of Acute Cardiovascular Compromise. AS-aortic stenosis; ASO-arterial switch operation; BAS-balloon atrial septostomy; BAV-balloon aortic valvuloplasty; ECMO-extracorporeal membrane oxygenation; IAS-interatrial stent; LPA-left pulmonary artery; CM-cardiomyopathy; PM-pacemaker; TAPVD-total anomalous pulmonary venous drainage; TOF/APV-Tetralogy of Fallot with absent pulmonary valve; TV-tricuspid valve

**Figure 3.**
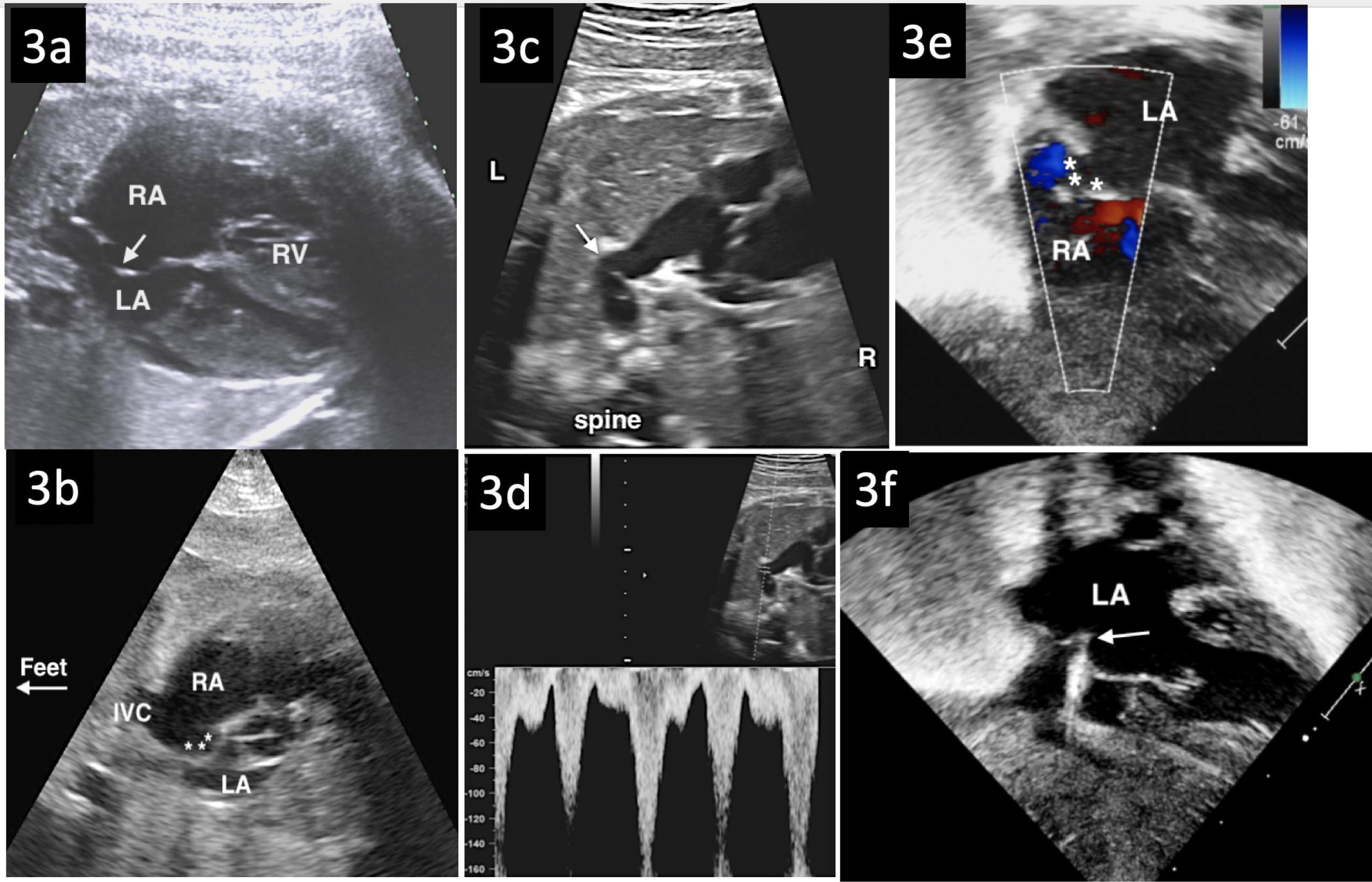
d-TGA/IVS with Restrictive Atrial Septum. (**A**) Cross-sectional 4-chamber and (**B**) sagittal (**B**) fetal echo images showing a tethered atrial septum with an aneurysmal foramen ovale flap (arrow A, * B) bowing to the left and tethered to the superior portion of septums ecumdum. An atrial communication could not be demonstrated. (**C**) 2D image of the ductus arteriosus demonstrating a discretely narrowed lumen (arrow). (**D**) Doppler interrogation of the ductus arteriosus showed increased systolic and diastolic velocities suggestive of ductal constriction. (**E)** Subcostal view at postnatal echocardiography revealed no demonstrable atrial communication, further suggested when (**F**) the septostomy catheter (arrow) could only tent and not cross the septum. With severe cyanosis and metabolic acidosis, the newborn underwent an arterial switch operation at <1 hour. LA-left atrium; RA-right atrium, RV-right ventricle.

**Figure 4.**
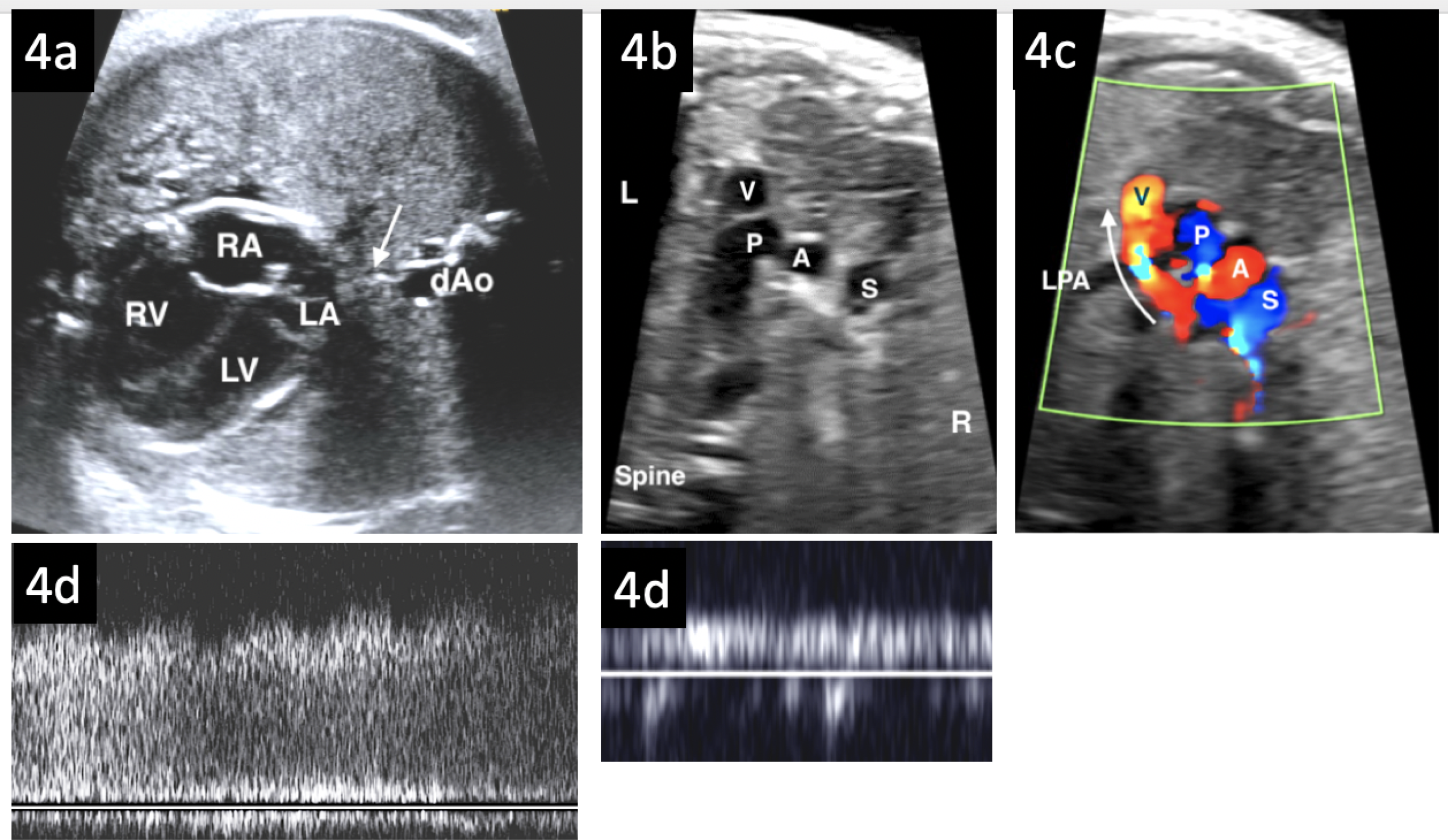
Supracardiac TAPVD with Obstruction. (**A**) 4-chamber image at 36 weeks showing right(RV) >left(LV) ventricular asymmetry and an increased distance (arrow) between the left atrium (LA) and descending aorta (dAo). Increased echogenicity of the lung parenchyma was suggestive of pulmonary edema. RA-right atrium. (**B**) In the 3-vessel view, the superior vena cava (S) and aorta (A) were of comparable in size, and there was a structure (V) to the left of the pulmonary artery (P). **(C**) By color Doppler the vertical vein (red flow and V) coursed over the left pulmonary artery (LPA) to join the left innominate vein. Where the vertical vein coursed over the LPA there was obstruction by color and (**D**) pulsed Doppler, with a mean gradient of 5mmHg. (**E**) Pulmonary vein flow was low-velocity and nonphasic flow in keeping with obstruction. This infant required ECMO at 4 hours for progressive cyanosis.

**Figure 5.**
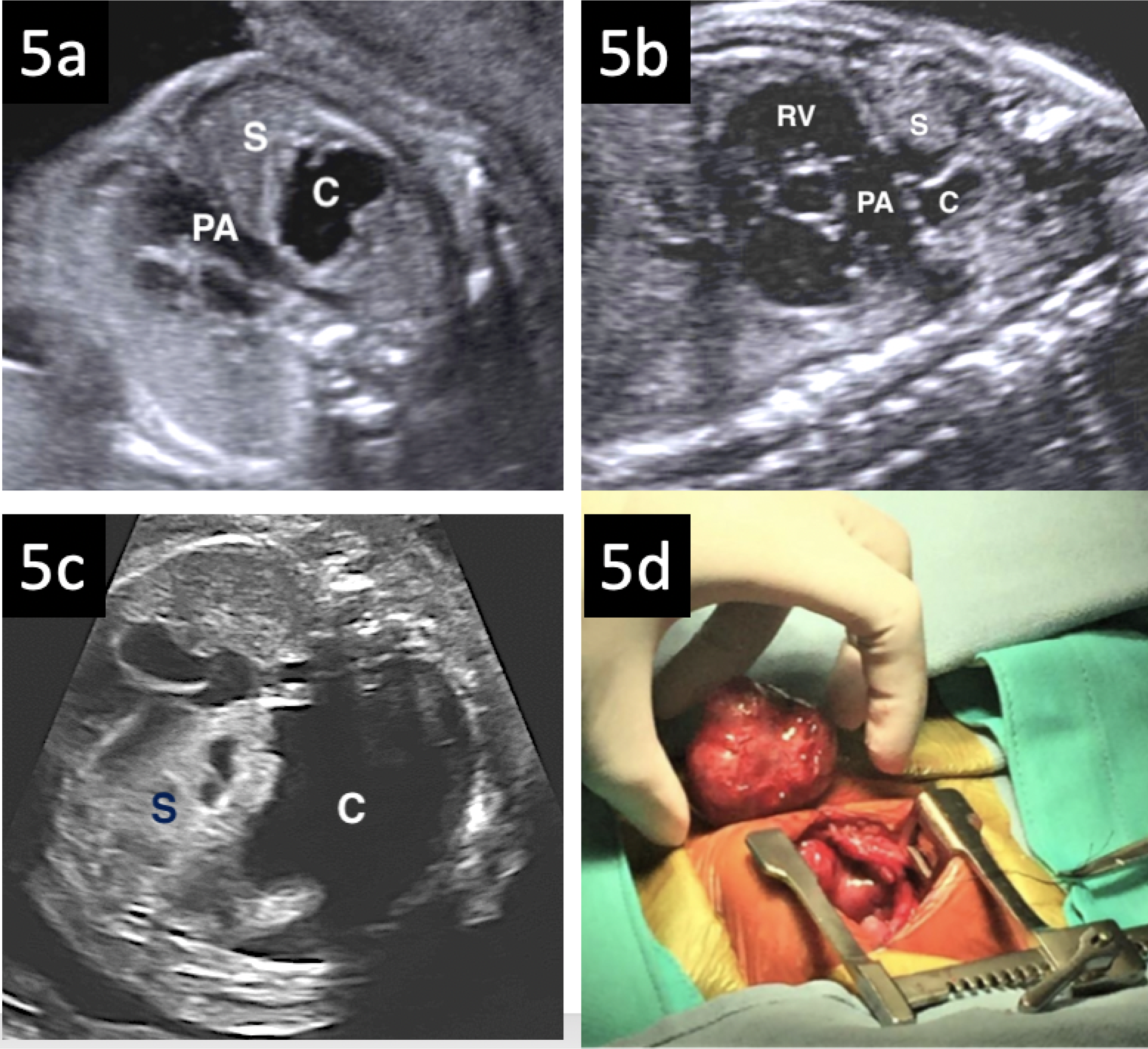
Cardiac Compression by a Large Mediastinal Tumor. **(A)** Cross-sectional and (**B**) sagittal fetal echo images at 23 weeks demonstrated a tumor in the superior mediastinum with cystic(C) and solid(S) components abutting the pulmonary artery (PA) and mild cardiac dextroposition. Rapid growth of the tumor from 30 weeks led to development of hydrops. **(C**) At 33 weeks the now massive tumor occupied much of the fetal chest with significant cardiac compression. These findings prompted preterm delivery. With progressive cardiorespiratory embarrassment, the tumor (**D**) was removed surgically 7 hours after birth. Pathology assessment confirmed a germ cell tumor, and an acute hemorrhage into the tumor was the cause of its rapid increase in size in utero.

Of the 40 fetuses, 33(82.5%) were delivered in the PCOR as planned at a median gestation of 37.4(range 32.7-39.9)weeks and median birth weight of 3.0(range 2.1-4.4)kg. Of these 33, 8 (24.2%) were delivered at <37 weeks’ gestation: 3 with twin gestations (2 dichorionic, 1 monochorionic at 35.0-36.0 weeks), 4 for evolving compromise/hydrops and/or non-reassuring obstetrical surveillance (32.7-36.7 weeks), 1 with spontaneous labor with rapid transport to the PCOR (33.6 weeks). Seven others of the 40 were delivered out of the PCOR at a median gestation of 38.4(range 37.4-40.0) weeks and median birth weight 3.3(range 2.8-3.7)kg, including 5 with d-TGA/IVS with early spontaneous labor and 2 with HLHS and RAS whose mothers declined a CS.

Overall, for the high-risk cohort, FE had a positive predictive value of 50% for predicting the combined outcome of intervention/ECMO/death at ≤2 hours and 70% within the first 24 hours. There was no mortality at delivery for those delivered in the PCOR, and survival to hospital discharge was 82%.

The thirty-three mothers (median age 31, range 19-36 years) with a PCOR CS delivery had spinal anesthesia. Eight had a known concomitant medical condition: 3 gestational diabetes, 1 chronic hepatitis B infection with cholestasis of pregnancy, 1 systemic lupus, 1 gestational hypertension, 1 asthma and 1 chronic kidney disease (G2A2 stage). The delivery represented a primary CS in the majority (28/33, 85%), including 13 primiparous mothers. All were successfully transferred back to the obstetrical hospital in stable condition within 3-5 hours of CS. Overall, there were no major maternal complications (e.g. postpartum hemorrhage, Caesarian hysterectomy, or infection), and total length of hospital stay for the mothers was a median of 2(2-4) days. Minor complications occurred in 5 following hospital discharge (1 seroma, 1 wound pain, 1 spinal headache, 1 mild dehydration, 1 mild superficial wound infection), and were treated as outpatients with mostly supportive treatment.

### d-TGA/IVS Delivery & Outcomes

Of 61 fetuses diagnosed with d-TGA/IVS during the study period, 15 were deemed high-risk (LOC4) and anticipated to require urgent balloon atrial septostomy (BAS) after birth. Ten of the 15 had a PCOR delivery (median gestation 38.6, 37.9-39.9weeks). Of these 10, 6 underwent urgent septostomy at ≤2 hours after birth for significant cyanosis despite intubation, medications and supplemental oxygen. In a 7^th^, an intact atrial septum necessitated emergent arterial switch operation (ASO) in the first hour. Two others with an atrial septal defect of <3mm underwent BAS at 6 and 9 hours, respectively, due to progressive hypoxemia. The 10th neonate with a redundant septum primum and 3-4mm interatrial communication, deemed challenging for BAS, underwent ASO at 32 hours post-delivery for hypoxemia.

With early spontaneous labor, 5 of the 15 d-TGA/IVS planned for PCOR delivery were born vaginally at the obstetrical hospital (median gestation 38.2, 37.6-38.7weeks). All 5 were transported to the Stollery Children’s Hospital: 4 underwent immediate successful BAS at ≤2 hours after birth for severe hypoxemia, and the 5th arrested on arrival requiring ECMO for stabilization. All 15 cases underwent ASO at a median age of 5(0-11) days, with all but one surviving to hospital discharge.

All 46 d-TGA/IVS cases classified as lower-risk (LOC 2/3) and encountered within the study period were delivered in our obstetrical hospital, 14(30%) by CS, and were safely transferred for specialized cardiac care. Six(13%), underwent successful BAS at ≤2 hours after birth for severe hypoxemia. An additional 9 underwent BAS at ≤24 hours (range 3-14 hours) after birth, and one preterm infant required a BAS on day 11. All underwent ASO and survived to hospital discharge.

For the entire cohort with d-TGA/IVS, FE had a sensitivity of 67%, specificity of 93%, and positive and negative predictive values of 80% and 87%, respectively, for predicting the outcome of cardiac intervention/ECMO/death at ≤2 hours after birth.

### HLHS Delivery & Outcomes

Of 52 fetuses diagnosed with HLHS during the study period, 7 were classified as high-risk for ACRI, requiring urgent atrial septoplasty. Five had PCOR deliveries at a median gestational age of 37(35-39)weeks and median birth weight 3.0 (2.5-4.0)kg. Three underwent atrial septoplasty with stenting at ≤2 hours of delivery. One of the 3, a twin, who delivered at 35 weeks with fetal MRI features of “nutmeg lung”, required ECMO after atrial septoplasty and demised at <24 due to disseminated intravascular coagulopathy with persistently high lactates and intraventricular hemorrhage. A 4th developed florid pulmonary congestion necessitating urgent atrial septoplasty at 6 hours, and a 5^th^ case with a redundant atrial septum, initially deemed not to require urgent atrial septoplasty, underwent early Norwood-Sano surgery at 36 hours given evolving hypoxemia and pulmonary venous congestion. Two of the 7 high-risk HLHS were delivered vaginally: one in a local hospital to a mother lost to follow-up from our service after the plan for PCOR delivery was made. This baby demised within minutes of delivery. Another mother who declined CS, had spontaneous labor and delivered vaginally at our obstetrical hospital. On arrival to our PC hospital, with profound hypoxemia the neonate required ECMO. Care was withdrawn at <24 hours due to severe end organ dysfunction. Of the surviving 4 of 7 high-risk HLHS infants, 3 underwent Norwood-Sano surgery at a median age of 5 (2 -9) days and 1 a hybrid procedure at 20 days. All four survived to hospital discharge. Survival of high-risk HLHS who had a PCOR delivery was 80% versus 0% without PCOR delivery.

All 45 low-risk fetal HLHS cases encountered within the study period were delivered at the obstetrical hospital, 16(36%) by CS, and were safely transferred for specialized cardiac treatment. None required intervention prior to Norwood-Sano or hybrid procedure and all survived to hospital discharge. For the entire cohort of fetuses with HLHS, FE had a sensitivity of 100%, specificity of 95%, and a positive and negative predictive values of 71% and 100%, respectively, in predicting the combined outcome of cardiac intervention/ECMO/death at ≤2 hours of delivery.

Other than the “low-risk” d-TGA/IVS cases requiring urgent BAS, no other prenatally diagnosed CD case encountered over the study period with intent-to-treat and not deemed at risk for ACRI required emergent intervention/ECMO or died at ≤2 hours after birth.

## DISCUSSION

A small but important subset of fetuses with major CD are at risk of ACRI shortly after birth and require specialized delivery planning to provide immediate stabilization with an intervention and/or medical treatment. With increasing experience in fetal CD diagnosis, the benchmark of a successful fetal cardiology program has shifted from diagnostic accuracy to predicting perinatal/neonatal presentation with detailed delivery care planning in an effort to optimize perinatal outcomes. Our results suggest that FE can be used to effectively predict most high-risk cases of d-TGA/IVS and HLHS with RAS, and the need for very early intervention in at least 50% of other cardiovascular pathology through a multidisciplinary team approach. We further describe a unique algorithm of an organized PCOR delivery remote from our quaternary obstetrical hospital, an approach to date that has received little attention.

Ideally, for the highest-risk fetal CD, delivery should occur in or within close proximity to the PC hospital to access the necessary infrastructure for resuscitation. Unfortunately, many programs internationally have a significant distance separating the delivery room from the PC hospital, making the ex-utero transport of a critically unstable neonate challenging. Even transporting such newborns from an adjoining building or between floors can, at times, be difficult. Alternatively, neonatal cardiac resuscitation can be orchestrated where the delivery occurs, but for many centers, this would be remote from other necessary equipment and personnel. Determining where delivery of high-risk cardiac newborns should occur is part of a complex decision-making process involving collaboration between multiple teams, and requires consideration of infrastructure, including facilities, equipment and personnel, within any program. It requires communication algorithms which take into consideration factors such as availability of expertise on site to offer both maternal and neonatal care, treatment options, the distance from the delivery room to the intensive care or interventional/surgical unit, and practical aspects of transporting a sick newborn safely and efficiently. An additional factor is availability of appropriate delivery and resuscitation space on the right day and time. The PCOR delivery approach used at our institution represents an important move towards minimizing cardiac mortality and morbidity of the highest risk fetal CD diagnoses by reducing crucial time to a life-saving intervention or stabilization. Arrival of the neonate in the intervention suite without prior significant cardiorespiratory compromise should have a positive impact on the success rate of intervention with a reduction in complications, and ultimately lead to improved short and long-term outcomes. Parents are key decision makers, and this type of perinatal care planning requires parental commitment (especially for the mother) to consent to this strategy of care.

Our study shows successful implementation of an algorithm of high acuity perinatal/neonatal care through a multidisciplinary, cardiac program-wide approach. We witnessed no important maternal morbidity beyond the CS, and mitigated mortality and morbidity risks for many affected newborns. We used a combined outcome of intervention/ECMO/death at ≤2 hours of delivery to evaluate our institutional experience of predicting such cases. The distance from our obstetrical to PC hospitals and the terrain covered during transport, including a heavily trafficked areas and dependence on a bridge, warranted this time cut-off for intervention. However, this only takes into consideration newborns requiring intervention; whereas, it did not capture risks of transporting a fragile newborn with potential for cardiopulmonary extremis and collapse in transit. Without a randomized trial, it is difficult to fully ascertain the benefits of avoiding transportation. However, the two cases requiring emergent atrial septoplasty, both arresting on arrival to our cardiac center necessitating ECMO and others requiring immediate intervention following PCOR delivery, emphasize the critical nature of such newborns and the need to balance risks of late prematurity as well as maternal CS outside of the obstetrical hospital versus, for some, potential arrest and even death. We attempted to mitigate maternal risks by only considering largely lower-risk mothers, moving the smaller but fully complemented obstetrical team for the procedure, and following a brief period of monitoring relocating the mother to the obstetrical hospital for ongoing postpartum care.

In a high-volume pediatric cardiac program, HLHS and d-TGA/IVS with RAS/IAS comprise the bulk of fetuses requiring cardiac interventions at birth as witnessed in our experience. It was reassuring that previously established FE-based parameters in retrospectively examined cohorts^8-13^ were fairly accurate in guiding delivery planning for most with these two lesions. For HLHS with RAS we found FE to be highly accurate (100% sensitivity and 95% specificity) in predicting postnatal pulmonary venous hypertension requiring decompression of the left atrium. Facilitated by improving prenatal detection of d-TGA in our province,^23^ we <u>prospectively</u> predicted a need for BAS at ≤2hours with a sensitivity of 67% and specificity of 93% and failed to predict the need for urgent BAS in only 13% of the others. As has been reported previously,^24^ there are still limitations in predicting d-TGA/IVS with RAS with resultant profound hypoxemia at birth, especially in late 3^rd^ trimester pregnancies. One advantage of our approach is that the infant with d-TGA/IVS is delivered at a time when personnel and equipment for BAS are immediately available for a day-time procedure, often performed during a short “honeymoon period” when the baby has not as yet become profoundly hypoxemic and acidotic as a consequence. Such interventions, under less duress, with earlier optimized mixing should contribute to less morbidity and mortality previously reported,^25^ and ultimately improved longer-term outcomes. Furthermore, immediate availability of other services, should a rare catastrophic event occur, provides further benefit of this approach.

For the other lesions in our cohort with limited published data to guide delivery planning, we formulated PCOR care plans based on anticipated pathophysiology and fragility in the perinatal transition with expert multidisciplinary consensus. With experience, we have modified our approach further, particularly for Ebstein anomaly and TOF/APV, including more routine use of fetal MRI to assess airways in the latter, where most infants may have been sufficiently stabilized for transport. For TAPVD, although we correctly predicted obstruction, we found it challenging to predict the exact timing of intervention. Low velocity non-pulsatile pulmonary vein flow aids in predicting obstruction, but in our cohort, one baby required acute intervention with pulmonary vein flow patterns more akin to RAS in HLHS. Another required ECMO by four hours and still another had surgical repair several hours after birth following necessary axial imaging. In the latter 2 cases, PCOR delivery provided additional time of relative stability for further imaging prior to repair. In a few cases we had missed opportunities and could have been more proactive in instituting the interventions without waiting for decompensation. Every institution has a learning curve when a new process is employed, and we would approach such cases more aggressively in the current era. Timely post-delivery review of individual cases has allowed us to continue to refine our algorithm.

### Limitations

The current study was a single-center experience over nearly a decade. The study was limited by its small sample size, particularly for more rare lesions, making it difficult to draw strong conclusions regarding the accuracy of FE for all CD. There was also a potential for intervention bias introduced by having a predetermined delivery and postnatal care plan, mitigated, to some extent, by relying on the newborn’s clinical status following the initial perinatal transition and a multidisciplinary team decision as to whether to proceed. Thus, if the baby could be stabilized, a planned early intervention did not occur. It is acknowledged that the transition norms of oxygen saturation, blood pressure and heart rates of many CD in the first 10-15 minutes after birth is an emerging area of study and that ongoing experience may alter our interpretation in the future regarding need for and timing of intervention.^26^ The study did not explore the potential long-term benefits of early restoration of adequate cardiac output and/or oxygenation, the impact on morbidity and mortality after further definitive surgical intervention or the longer-term impact, including on neurodevelopment. Additionally, the limited number of mothers undergoing CS in the PCOR may not truly reflect the potential short- and long-term implications of obstetrical surgery performed in a children’s hospital.

## CONCLUSION

FE predicted need for urgent intervention in majority with d-TGA/IVS and HLHS, and in half of the entire spectrum considered high-risk. Based on this experience, FE can be used to predict ACRI, especially for d-TGA/IVS and HLHS with RAS, thus allowing for planned perinatal and acute neonatal specialty delivery room care. Delivery-planning needs to be tailored to a given institution’s geographical factors and available infrastructure to take full advantage of the prenatal diagnosis. For some, such as our own, a well-organized algorithm which includes delivery within the PCOR of the highest acuity fetal CD cases may be warranted and can be successfully implemented with multidisciplinary involvement for the care of the mother and baby.

## Data Availability

All salient data is available in the manuscript

## Non-standard Abbreviations and Acronyms

ACRI: acute cardiorespiratory instability
ASO: arterial switch operation
BAS: balloon atrial septostomy
CD: cardiac disease
CS: Cesarean section
d-TGA/IVS: dtranspoisiotn of the great arteries/intact ventricular septum
ECMO: extracorporeal membrane oxygenation
FE: fetal echocardiography
HLHS: hypoplastic left heart syndrome
LOC: level of care
PCOR: pediatric cardiac operating room
RAS: estrictive atrial septum
TAPVD: total anomalous pulmonary venous drainage
TOF/APV: tetralogy of Fallor with absent pulmonary valve syndrome

## Acknowledgements

We want to acknowledge the critical contributions of our maternal-fetal-medicine nurses, operating room and obstetrical managers as well as other staff who have made the execution of these deliveries possible.

